# The Forgotten Hip Fracture: Outcomes Of Lower Limb Periprosthetic Fractures

**DOI:** 10.1101/2022.10.28.22281643

**Authors:** Manu Shrivastava, Vinci Naruka, Stephen McDonnell

**Affiliations:** Department of Orthopaedics and Geriatrics, Cambridge University Hospitals, Hills Road, Cambridge, CB2 0QQ

## Abstract

**Introduction:** There is an increasing prevalence of primary hip fractures and peri-prosthetic hip and knee fractures. There is uncertainty about how best to manage peri-prosthetic fractures, and they do not attract the same financial incentives and management guidelines.

**Methods:** A retrospective review of medical records was performed at a large academic teaching hospital between October 2014 and September 2016. Seventy-three patients who sustained periprosthetic fractures of the hip and knee were identified. These were compared with outcomes for the hospital recorded on the National Hip Fracture Database.

**Results:** There were difference in the baseline characteristics between the two groups, with PPF patients having a younger age, lower frailty score and being more likely to be female. There were lower rates of pre-operative assessments for the PPF group. Given the differences in baseline characteristics, their post-operative scores could not be reliably statistically compared.

**Discussion:** Patients with PPFs may have different characteristics and outcomes to patients presenting with primary hip and knee fractures. More work is needed to better characterise this patient group.

**Conclusion:** patients sustaining periprosthetic fractures represent a distinct patient group to those with primary hip fractures.

## Introduction

The increasing healthcare burden of primary hip fractures is well known. They occur in 0.1% of the population but are likely to increase in prevalence with the UK’s ageing population ^1^. Due to the significant mortality and morbidity that results from these fractures, there has been a concerted effort to improve outcomes. The Best Practice Tariff (BPT) for primary femoral neck fractures has led to a significant improvement in outcomes since its introduction. Mortality rate has dropped from 10.9% to 7.1% in just 8 years ^1^.

Whilst still uncommon, peri-prosthetic fractures are increasing in prevalence in line with arthroplasties. Their current prevalence has not been established due to a lack of long term follow up, but a 20-year probability may be as high as 11% ^2^. Whilst figures on incidence vary, it is thought that sustaining a primary hip fracture increases the relative risk of sustaining a further fracture compared with the normal population - one study found this risk to be 45-fold in females aged 55-64 years ^3^].

Peri-prosthetic fractures (PPFs) have a devastating impact on health. They are different to primary fractures in terms of the demographic that is affected, the management, and the outcomes. They have a high mortality rate, ranging from 11-18% at one year ^4–6^. This is similar, or even slightly higher, than equivalent rates for primary fractures. Moreover, there is substantial morbidity, and a poor functional outcome after PPF surgery ^5,7^.

Despite the significant and increasing impact they have both on health and on healthcare, there is no guidance on ‘Best Practice’ how best to manage these patients. The financial incentive attached to the BPT has encouraged optimal management and implementation of guidelines for primary hip fractures. Whilst a specific incentive targeted at PPFs does not exist, the cost incurred by suboptimal management can be substantial. PPFs are thought to cost up to £10,000 per patient, with the bulk of the cost related to the length of stay ^8^.

The purpose of this study is to compare the pre-operative assessment and post-operative results of peri-prosthetic hip and knee fractures with primary hip fractures. Specifically, factors that are thought to be related to worse outcomes - frailty, post-operative complications, and delirium - are analysed in more depth.

## Materials and Methods

A retrospective review of medical records was performed on patients who presented with peri-prosthetic lower limb (knee and hip) fractures at a large academic teaching hospital between October 2014 and September 2016. Local clinical governance and ethical standards were followed. Seventy-three patients were identified as eligible for analysis, and their records were interrogated using an electronic patient record system.

Information was collected concerning epidemiological characteristics, orthogeriatric risk assessments, type of fracture and procedure, and outcome. The raw data for this project are available on request.

The abbreviated mental test score was used to assess cognitive state. The Rockwood clinical frailty score was used to grade frailty ^9^. The American Society of Anaesthesiologists (ASA) grade was used pre-operatively ^10,11^. Delirium was identified using a 4AT score ^12^.

Comparisons were made with primary hip fractures treated at Cambridge University Hospital (CUH), using data from the annual National Hip Fracture Database Report ^1^.

Information in the NHFD is collected yearly, but does not contain specific assessments such as frailty or delirium, as this data was not routinely collected until September 2016. Therefore, a smaller group of patients was selected for in-depth analysis; these patients had sustained primary hip fractures between September 2016 - February 2017.

Standard summary statistics were used throughout. Independent t test was calculated for statistical significance. There were no corrections for multiple testing.

## Results

Admission statistics were calculated for the peri-prosthetic fracture and compared to results from primary hip fractures managed at our hospital on the National Hip Fracture Database ^1^. The results are shown in figure [as].

Patients with peri-prosthetic fractures were found to be younger (78 vs 83 years, p=0.001), more likely to be female (p=0.03), and less frail (4.4 vs. 5.0, p=0.03) than patients with primary hip fractures. Given these differences in baseline characteristics, their post-operative outcomes could not be reliably compared.

Given the different characteristics of each population, further analysis was performed to compare management. The levels of pre-operative assessments done in the PPF group were consistently and significantly lower than for primary neck of femur fractures, as shown in figure 3.

**Figure 1:**
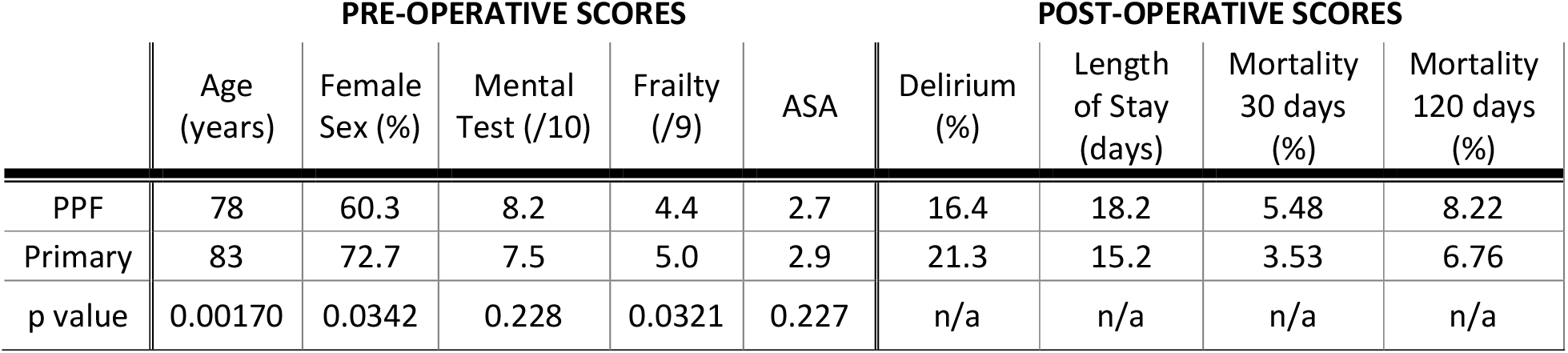
summary table demonstrating baseline characteristics and clinical outcomes of patients with periprosthetic fractures (PPF) compared to primary hip fracture outcomes at the trust. All figures given represent the mean average for each parameter. The abbreviated mental test score was used to assess cognitive state. The Rockwood clinical frailty score was used to grade frailty. The American Society of Anaesthesiologists (ASA) grade was used. Delirium was identified using a 4AT score. Given that the two groups had different pre-operative baseline scores, further comparisons were not calculated for post-operative scores.

**Figure 2:**
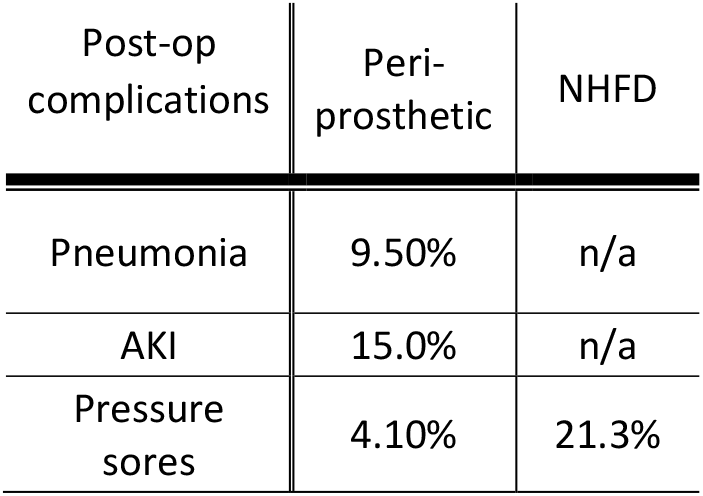
comparison table of post-operative complications seen in peri-prosthetic hip and knee fractures compared to figures for primary hip fractures. n/a represents missing data from the NHFD. AKI stands for acute kidney injury.

**Figure 3:**
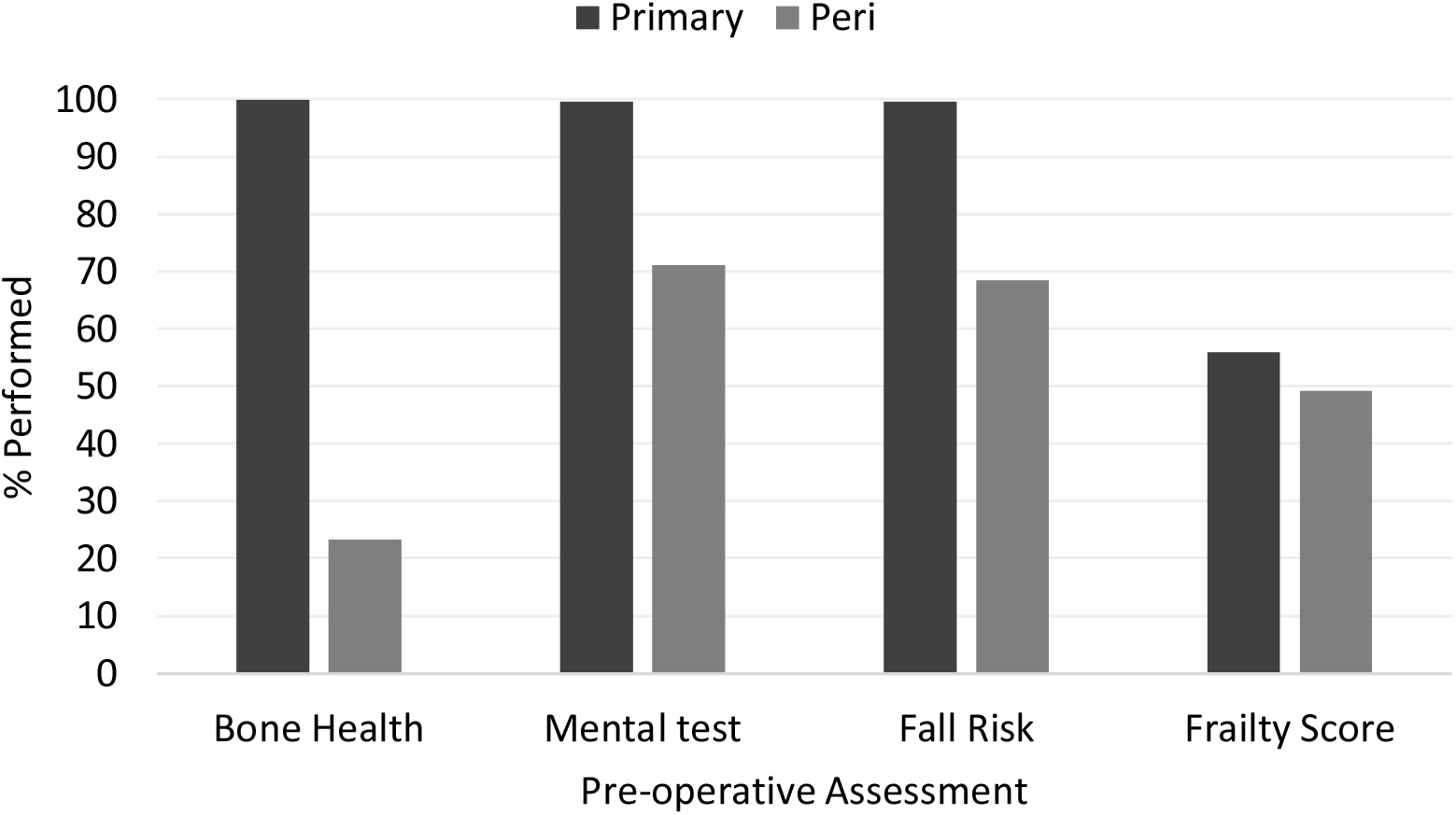
bar chart comparing the percentage of completed assessments for primary hip fractures (NHFD) and periprosthetic fractures at the hospital.

The type of operation performed after a peri-prosthetic fracture varies between hospitals and between surgeons. The type of fracture sustained and the management was analysed. Of the periprosthetic fractures analysed, 68.5% were hip, and 31.5% were knee fractures.

Most fractures (62%) were B1 or B2. Open reduction and internal fixation was used for 75% of fractures.

## Discussion

Patients who have sustained peri-prosthetic fractures represent a distinct group to those with primary hip fractures. This study found differences with regards to their age and frailty. It is difficult to draw definitive conclusions between the two groups based on this study alone given their varying baseline characteristics.

In comparison to the reported NHFD for primary fractures, there is a lower pre-operative assessment rate conducted for PPFs. It should be noted that PPFs include both hip and knee fractures. To pre-empt any avoidable bias, all admissions reports and documentation was analysed and followed up to the pre-operative date. In addition, pre-operative assessments for PPFs are carried out by a team of two orthogeriatricians who perform all the relevant investigations before an orthopaedic review.

Looking at the data, it is possible to clarify some of the lower values. The bone health is assessed by a DEXA scan, which is normally carried out post-fracture. However, if primary hip surgery and a DEXA scan had been performed recently, another was not deemed necessary to repeat.

Furthermore following the recent osteoporosis national guidelines, in patients aged 75 years or over, a DEXA scan may not be required if the responsible clinician considered it to be clinically inappropriate or unfeasible ^14^

In addition, many of those who did not have these assessments performed were transfers from other hospitals (data not shown). Of the ones that were assessed, peri-prosthetic fracture patients were found to have a worse mental state, but a lower level of frailty.

There is no specific guidance on how to manage to manage this group of patients. Primary hip fracture results have improved with the introduction of the NOF Tariff ^1^ which stipulates certain pre-operative assessments and time limits for operations. By contrast, our results show that there are lower rates of specific pre-operative assessments for those with PPFs (figure 3). One reason is that many of PPF patients are transfers from other hospitals to CUH, a tertiary referral centre. Many of the assessments are done in the original hospital, and so are not documented in CUH records. It remains to be established which assessments are necessary and accurate in reflecting the outcome of these fractures.

The lack of guidelines for best practice perhaps stems from the complexity and large variation in PPF patients and in their comorbidities. It has resulted in multiple approaches in the management of peri-prosthetic fractures, resulting in a large variation pre-operative assessments and in outcome. Optimum management will undoubtedly have a significant impact on the quality of life for these patients.

The type of surgery performed is naturally very different for PPFs. It may be longer and require more experienced surgeons, which perhaps delays time to theatres. The types of fracture and operation performed are demonstrated in figures 4 and 5. This is consistent with other studies ^2,5,15,16^. Further, the complexity of a PPF and the difficulty in deciding the best management option can potentially result in a longer length of stay, which additionally poses risks of developing hospital-acquired infections and complications.

**Figure 4:**
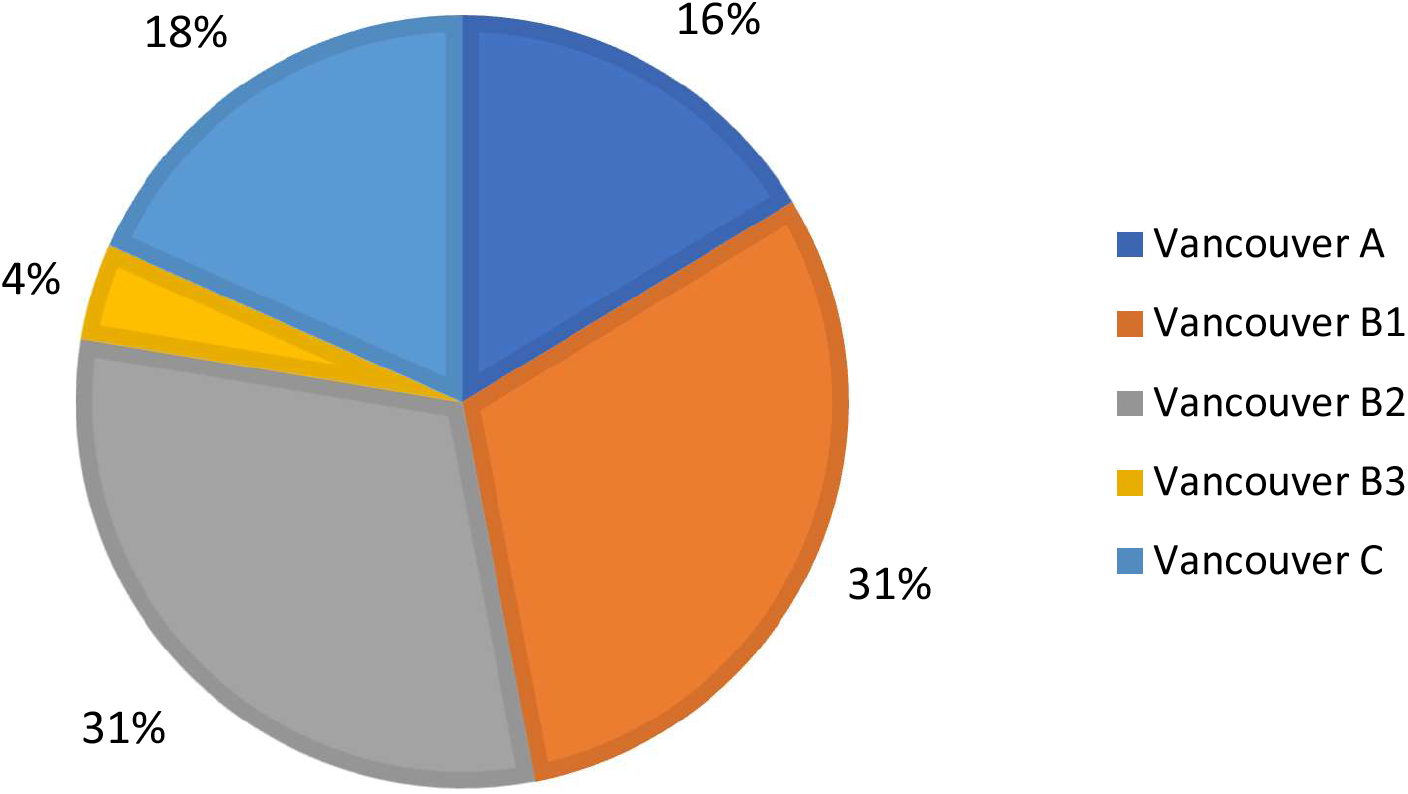
shows the type of periprosthetic fracture sustained classified by the Vancouver classification ^13^.

**Figure 5.**
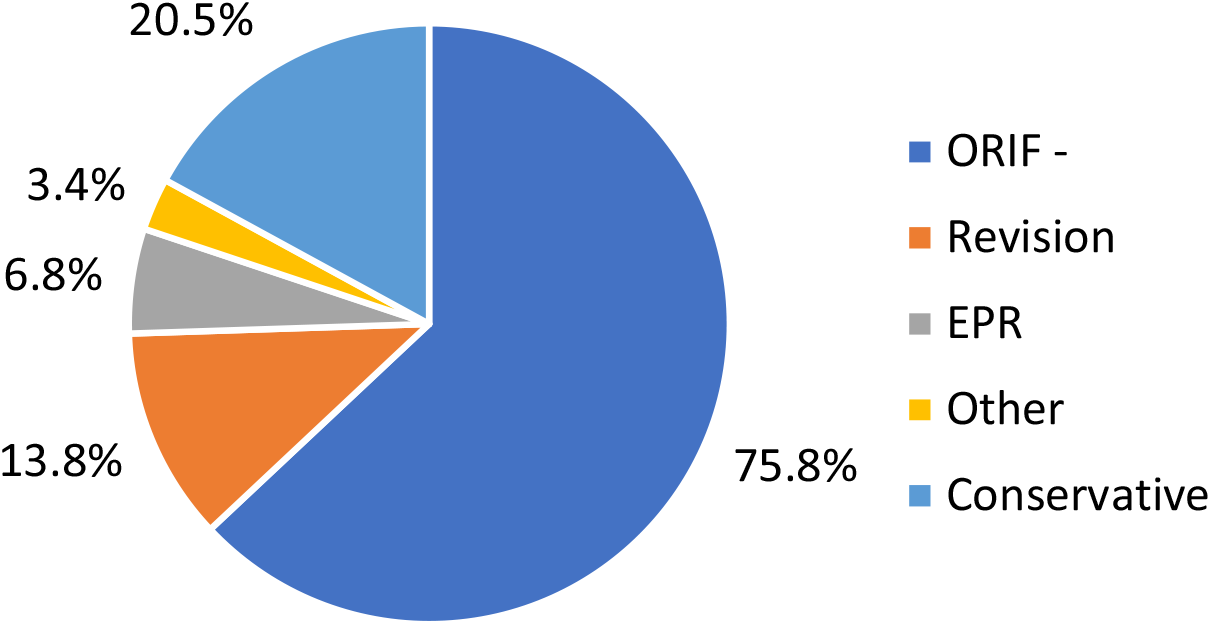
shows management strategies used. Twenty percent of fractures were managed conservatively; the rest underwent surgical management with implants/techniques displayed above. ORIF stands for open reduction and internal fixation (ORIF). EPR stands for endoprosthetic replacement.

The Vancouver classification distinguishes between different types of PPF. It is based on the anatomy of the fracture in relation to the prosthesis, and has been used to stratify management and prognostication. Whilst its usefulness is limited clinically ^5^ and stratification has not been used in this study, data are provided to show that surgical management of this cohort of patients is comparable to other centres.

There is a financial incentive attached to the NOF tariff and it is profitable for trusts to prioritise these procedures. Hip fractures nationally cost £1 billion, most of which is due to the increased length of hospital stay of these patients ^8,17^. The average length of stay may be longer for PPF patients and the healthcare cost may be considerable.

There is a need for more guidance on the management of PPFs. Increasing resources and medical input may be warranted, as well as an increase in availability of senior surgeon cover.

## Conclusion

Peri-prosthetic fractures have different baseline characteristics and lower reported pre-operative assessment rates compared to primary hip fractures in the population studied. There is a need to increase awareness of this group, and to implement specific pathways for their care. More studies and guidelines on this area would be worthwhile.

## Data Availability

All data produced in the present study are available upon reasonable request to the authors.

